# Gaming the peer review system: a sophisticated review mill in medicine highlights the need to ensure reviewer integrity

**DOI:** 10.1101/2025.10.20.25338343

**Authors:** M. Ángeles Oviedo-García, René Aquarius, Dorothy V. M. Bishop

## Abstract

**Background:** A review mill is a network of researchers who game the peer review system to apparently boost their citations. Members write generic review reports containing suggestions for citations to the work of those in the review mill. Here we report compelling evidence for a review mill in the field of gynecologic oncology.

**Methods:** The first example of a peer-review report from a review mill was observed by chance. It contained ‘boilerplate’ comments, such as: “Methodology is accurate and conclusions are supported by the data analysis” as well as suggestions that specific PubMed IDs be cited. We searched the internet using Google for review reports using the same boilerplate. We coded all review text to quantify similarities between review reports and compiled a list of citations suggested by reviewers. For 59 of 119 nonanonymous reviews in this target set, we identified a second review of the same article to act as a comparison.

**Findings:** We identified a set of 195 review mill reports that shared verbatim or highly similar boilerplate text from 170 targeted articles. 186 reports suggested citing at least one article which was co-authored by the reviewer or another member of the mill. Authors of 142 articles complied with some or all suggestions for citation. Nine of the reviewers in this review mill contributed 5 or more reviews; four had acted as editors for articles in the target set, and five had prolific peer review histories on Web of Science. Boilerplate text and self-citation recommendations were rare in the comparison reports.

**Interpretation:** Review mills threaten both the scientific record and patient safety when clinically relevant articles are improperly scrutinized during peer-review. We recommend that publishers adopt open peer review and transparently report the editors responsible for handling papers, to make it easier to detect review mills.

## Introduction

Editors and peer reviewers play a crucial role as gate-keepers of the scientific literature (1) but in recent years, concerns have been raised about so-called ‘review mills’ infiltrating the publishing system (2,3). Review mills use reviewers with established credentials who write boilerplate reviews containing generic and vague recommendations that can be tracked across different peer review reports. They frequently include coercive citation requests that boost citations to the work of the reviewer or to others in the reviewer’s network.

Review mills are typically unidentifiable unless peer review reports are made public, so that commonalities between review reports can be identified. Even so, they are not easy to track down, because they require in-depth scrutiny of reviews. Few review mills have been documented: those that have been described may include nonsensical text (4), which can make detection easier, and the review millers have typically not been based in Western countries.

In this report we describe a review mill run by well-established, Italian physicians in the fields of gynecology and oncology, mostly affecting papers with clinical implications; this is an alarming development as it means that the quality of research literature in this area may be damaged because peer review has been compromised.

We report here results of an exploratory investigation documenting the methods we used to study the review mill, explaining how we established similarity between reports that showed beyond reasonable doubt that peer reviewers were working together. Our goal was to develop an analytic approach that would throw light on how review mills operate, and to compare reviews from the review mill to a comparison set of reviews from other reviewers of the same articles.

## Methods

A chance observation unearthed the first example, which used what we term ‘boilerplate’ terminology: sentences or phrases that are generic, insofar as they do not refer to content that is specific to the article, and which are found across reviews of different articles. Examples of boilerplate responses are:

- *“In my honest opinion, the topic is interesting enough to attract the readers’ attention*.*”*
- *“The authors have not adequately highlighted the strengths and limitations of their study. I suggest better specifying these points*.*”*

Subsequent review reports were identified by a snowball search, where we extended the collection of search terms as we found additional instances of boilerplate terms in reviews. We used three types of Boolean Google (search operator) searches: “reviewer” AND

a. A sentence or word chain from a report that met our criteria, i.e. what we term “boilerplate” wording AND/OR
b. PMID numbers from a report that met our criteria (see Table 1) AND/OR
c. Reviewers’ names from a report that met our criteria (see Figure 1)

**Table 1:** Numbers of articles included in the target set, by publisher and journal*.

| Publisher/journal | N articles |
| --- | --- |
| MDPI |  |
| Cancers | 16 |
| Diagnostics | 15 |
| International Journal of Molecular Sciences | 14 |
| Current Oncology | 5 |
| Biomedicines | 5 |
| Other | 20 |
| PLOS ONE | 36 |
| BMC |  |
| BMC Women's Health | 12 |
| BMC: Other | 4 |
| AME Publishing Company |  |
| Translational Cancer Research | 9 |
| Gynecology and Pelvic Medicine | 3 |
| AME: Other | 3 |
| BMJ Open | 11 |
| Qeios | 7 |
| F1000 Research | 4 |
| Baishideng Publishing Group: World Journal of Clinical Cases | 3 |
| Sage | 2 |

\*Individual journals shown for cases with 3 or more articles in the target set

**Figure 1.**
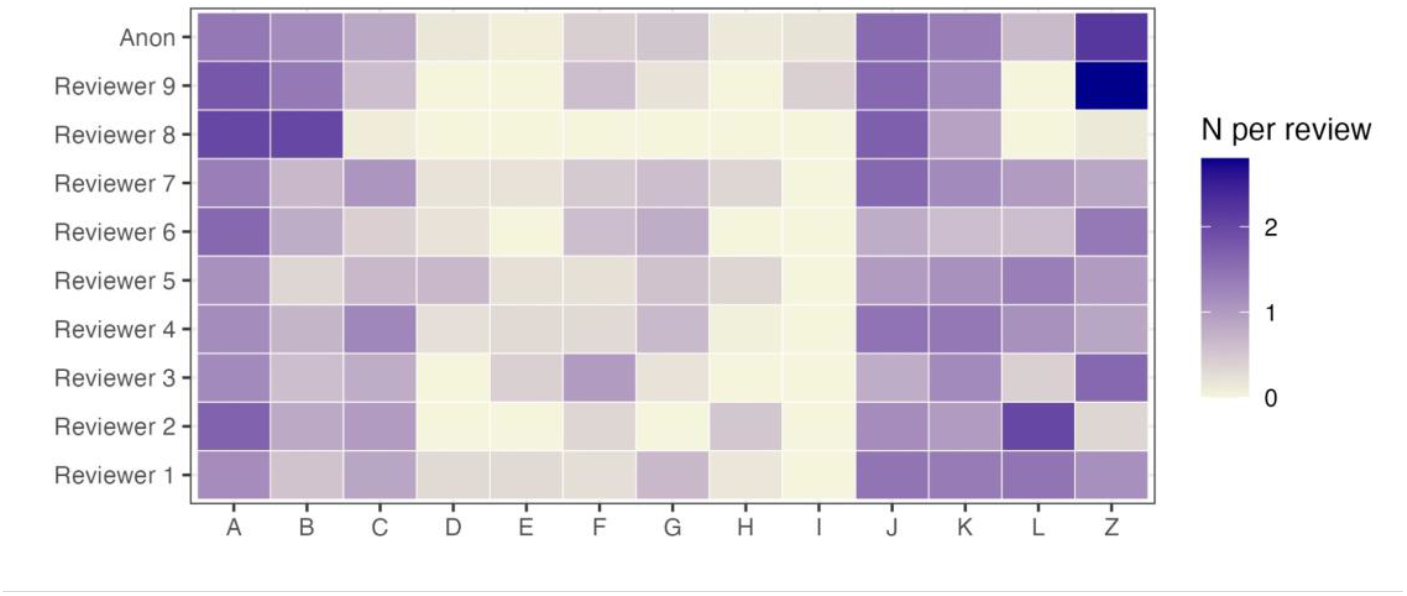
Heatmap where darkness of colour indicates average number of comments of that type per review by each of 9 reviewers, and by anonymous reviewers (Anon). For key to codes see Table 2.

Search results found between 7th November 2024 and September 1st 2025 were retained as ‘review mill reports’ when the report contained at least one (almost) verbatim sentence. The set of review mill reports was archived using the Wayback Machine (http://wayback.archive.org/). All the articles that were peer reviewed by the review mill will be referred to as the ‘target set’.

**Table 2.**
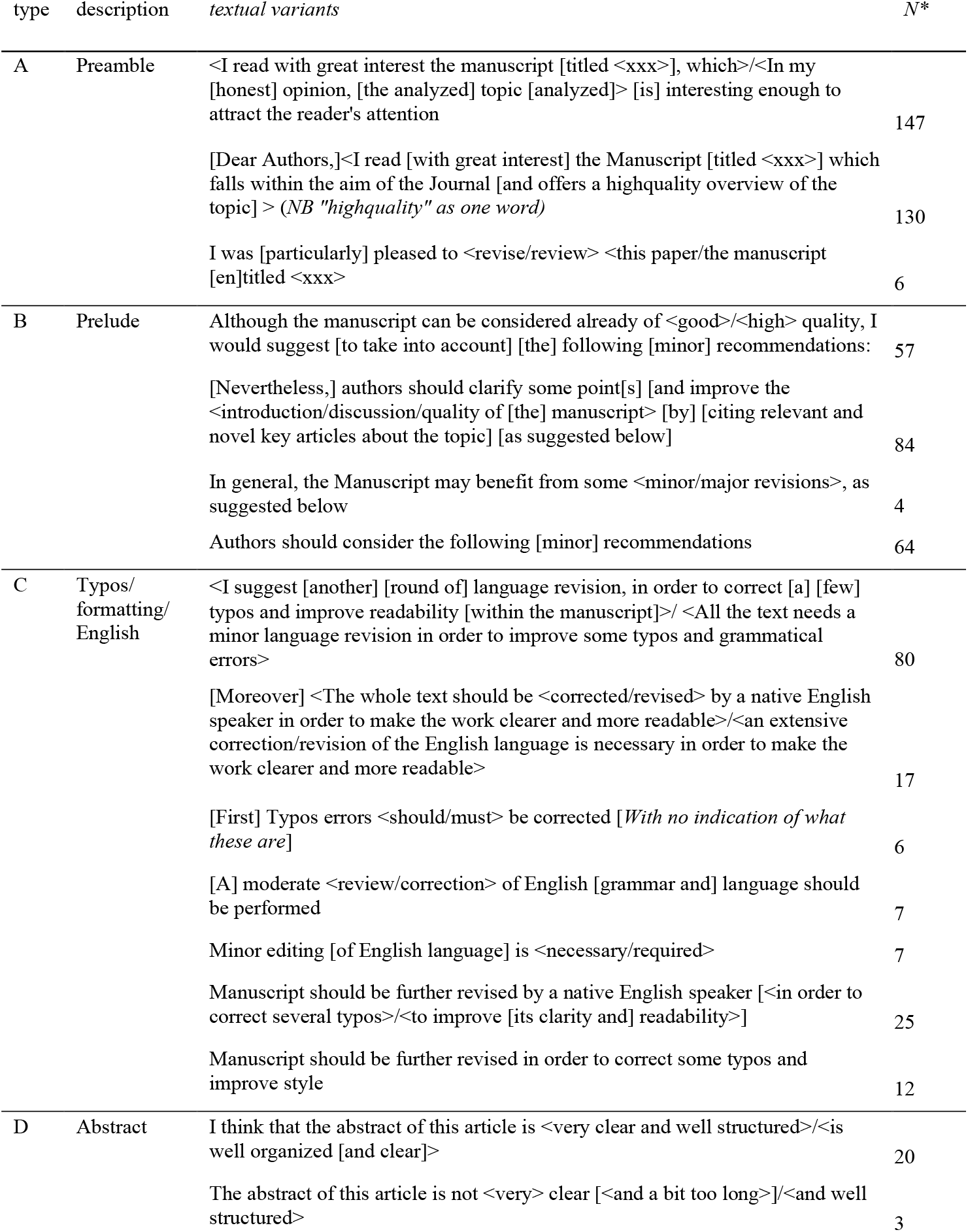
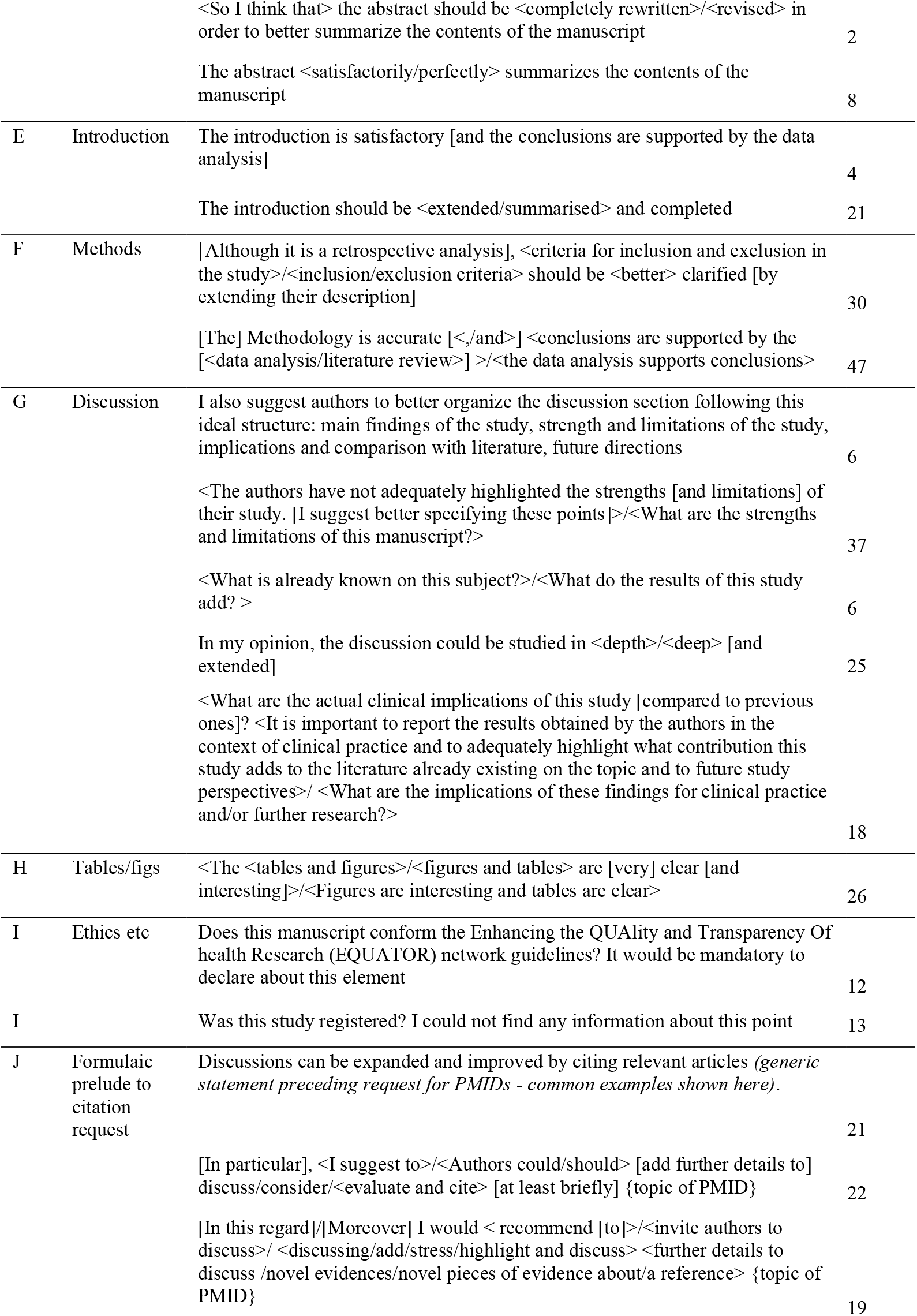
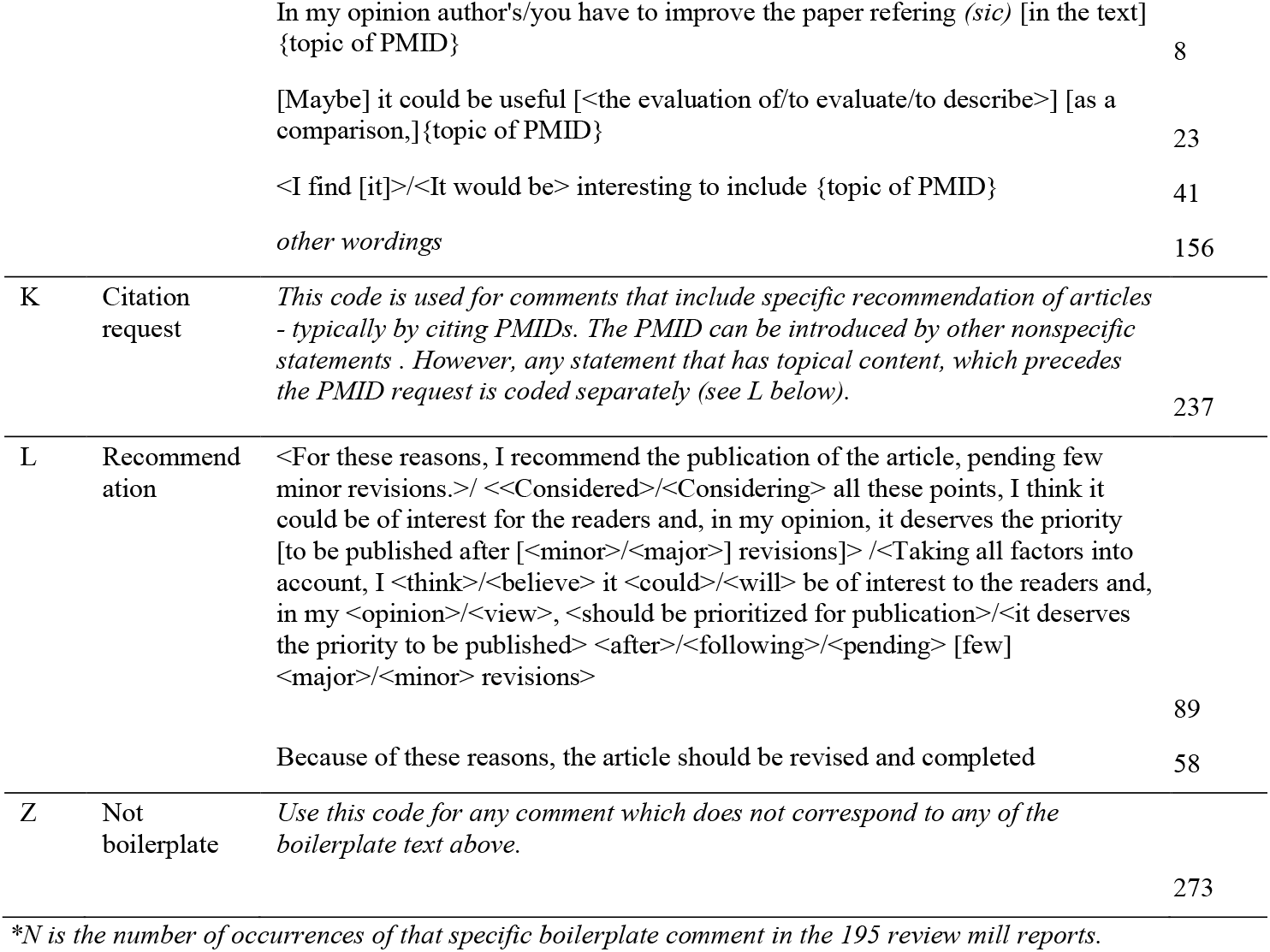
Coding scheme for boilerplate language in reviews. Italicised text gives some explanation. Optional phrases in square brackets; alternate wordings in pointed brackets separated by slash.

We also performed an identity check on reviewers, using links from open peer reviews to a valid ORCID or the reviewer’s full name and recorded their review history and H-index from Web of Science between September 30th and October 7th, 2025.

The whole set of boilerplate terms was scrutinised to create a coding framework, described more fully below. This was developed iteratively by identifying common sentences and word chains and grouping together those with closely similar wording. The three authors resolved by discussion how to handle cases where comments were closely similar but not identical. A separate code was used for reviewer comments that did not match our definitions of boilerplate.

Each review mill report was given an ID code, and all reviewer comments were then coded using the same framework. In addition, the PMIDs or other requested citations (DOIs or titles) were listed for each review.

We cross-tabulated the occurrence of boilerplate terms against the reviewer name to create a table showing the number of instances of that term being used. A similar analysis was done to cross-tabulate PMIDs mentioned in the review report against the name of the reviewer.

Reviewers are identified by numbers in this report.

A comparison group of 59 review reports was selected by focusing on the nine review millers who produced the most review reports, and selecting 50% of articles they had reviewed to provide a comparison review meeting these criteria:

a. consisting of at least three sentences;
b. the reviewer was not included in the review mill group; and
c. if possible the review was by a named reviewer (though an anonymous review was used if no other review met criteria a and b).

Any suggested citations in this comparison set of reviews were checked to see if they involved articles authored by the reviewer.

The comparison set of reviews allowed us to address three questions. First, are boilerplate comments a common finding, perhaps derived from language from instructions to reviewers? Second, is the number of non-boilerplate comments unusually low in reviews that contain boilerplate? Third, are suggested citations to the reviewer’s work unusually common in reviews with boilerplate text?

## Results

We identified 195 review reports from the peer review mill, which shared verbatim or highly similar boilerplate text with other publicly available review reports. The review mill reports belonged to 170 articles published between February 6th 2019 and July 7th 2025. This target set included 19 articles with two review-mill reports, and three articles with three review-mill reports. All but one of these cases involved different members of the review mill reviewing the same article. In the other case, three reviews by the same reviewer of the same article were identified on our search corresponding to three rounds of review; this reviewer barely modified their review report other than to suggest new references to be cited. Details of articles with review mill reports are available online as Supplementary Data 1.

Of the 195 review-mill reviews, 186 suggested at least one article for citation co-authored by the reviewer or another member of the review mill. In most cases, the request for citation did not identify the authors of the recommended articles, but rather referred to them by PubMed ID number (PMID), Digital Object Identifier (DOI), or title; only 21 review reports gave any information about authors when suggesting a citation. The median number of requested citations per review was two (100 reviews), with a range from 1 to 12.

The full set of coded peer review comments is available online (Supplementary Data 2).

### Journals/publishers

We identified several publishers and platforms that have been targeted by the review mill. These were MDPI, PLOS, BioMed Central, AME Publishing Company, BMJ Publishing Group, Qeios, and F1000 Research (see Table 1). Note that we can detect reports from the review mill only for journals that adopt open peer review practices.

### Cross-tabulation of reviewers with boilerplate comments

Table 2 shows the boilerplate codes and numbers of occurrences of different wordings. To simplify data presentation, codes are grouped into 13 categories.

Figure 1 is a heat map where darkness of the colour shows the mean number of instances of the review codes for each of nine reviewers and for anonymous reviews. The underlying data are available online (Supplementary Data 3).

Note that the more common boilerplate codes were used by all the reviewers. The final column (Z) shows non-boilerplate comments.

### Cross-tabulation of reviewers with suggested PMIDs

We use the term “coercive citation” for comments where a member of the review mill suggests that the article should be revised to contain mention of an article by themselves or another member of the review mill. There were 151 individual articles suggested as coercive citations, with many articles suggested several times, including by different reviewers. The heat map in Figure 2 shows the number of coercive citations, and the superimposed asterisks show whether or not the reviewer was an author of the coerced article. Underlying data are available online (Supplementary Data 4).

**Figure 2.**
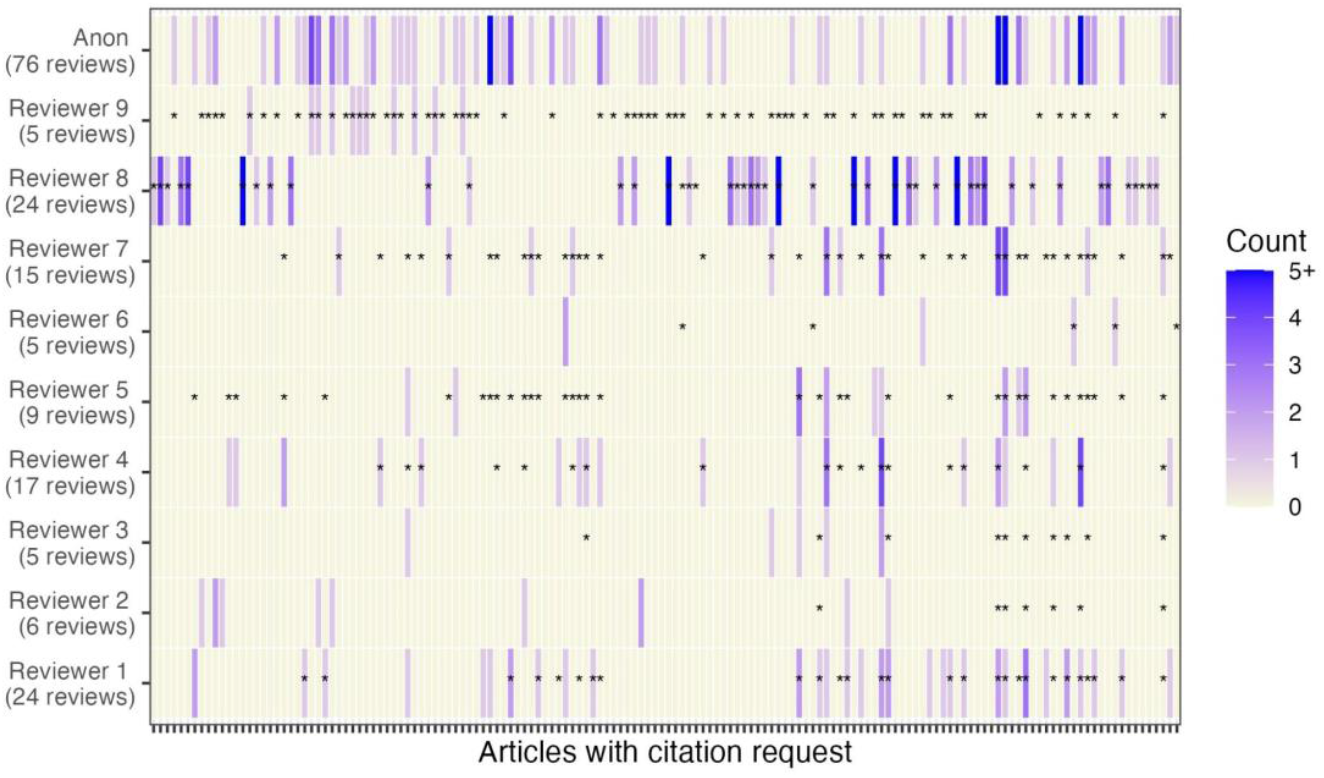
Heatmap where the x-axis shows a line for each of the 151 articles that reviewers suggested, and the depth of colour indicates the number of reviews where that article was suggested. Asterisks denote articles authored by the reviewer. The table containing full data used to create heatmap, including article PMIDs, is available online as Supplementary Data 4.

We also noted that when more than one review miller was involved in reviewing an article, they typically suggested a complementary set of references for citation (see Supplementary data 1 for examples).

We also noted for each PMID, how often a reviewer suggestion for a citation was successful (see Supplementary data 4). For 14 review reports this could not be evaluated, because there was no indication that the article had been revised after peer review: these were articles on platforms that adopted a “preprint, review, publish” model, i.e., Qeios and F1000. Of the remaining reviews, the final article was scrutinised to check if any of the suggested citations had been incorporated in the references. 142 (78%) of review reports led to inclusion of all or some of the suggested references in the article; in the remaining 39 (21%) authors either challenged the suggestion (e.g. by stating the reference was not relevant) or ignored it.

### Comparison reviews

The first question addressed by analysis of comparison reviews was whether boilerplate comments may arise from reviewers following journal instructions to reviewers. If so, we should expect to see them in comparison reviews.

The comparison and review mill reports were similar in number of coded statements; comparison set mean = 10.7 statements (SD = 5.81), review mill set mean = 9.8 statements (SD = 3.86). There were only 16 cases of boilerplate in 633 coded comments in the comparison reviews, compared with 1604 boilerplate coded comments out of 1875 coded comments in the review mill reports. Furthermore, five boilerplate codes in the comparison set came from the same review report. A check of that reviewer’s publications indicated she had coauthored articles with a member of the review mill. This was also true of another comparison reviewer who had suggested their own article for citation.

We considered whether the review mill reports might include non-boilerplate content in addition to the boilerplate. This was generally not the case: the mean number of non-boilerplate comments for the 59 comparison review reports was 10.4 (SD = 5.79), whereas for the 195 review mill reports it was 1.4 (SD = 3.15).

A further question was whether suggested citations to the reviewer’s work are unusually frequent in the review mill reports. After all, there are circumstances when it is reasonable to suggest that authors cite the reviewer’s work. There were only four instances of this in the comparison set. Suggested citations to work other than that authored by the reviewer were rare in both groups; there were 10 review reports including such requests in the comparison set, compared to 14 in the review mill set. The latter typically co-occurred with suggestions to cite the reviewer’s work.

### Academic editors of articles in the target set

Identity of academic editors was listed for 92 out of 170 articles. In 20 cases the academic editor was part of the review mill (three cases Reviewer 5, five cases Reviewer 7, two cases Reviewer 8, and ten cases Reviewer 9).

### Characteristics of members of the review mill

All members of the review mill were medical doctors with hospital and academic affiliations in Italy. They varied in productivity, as shown in Table 3. Information in this Table was largely compiled from Web of Science, supplemented by other sources such as ORCID and SciProfiles.

**Table 3:** Background information about members of review mill. All information was obtained from Web of Science, ORCID, or SciProfiles.

| ID | H-index | Awards | Editorial Board Memberships | Verified Peer Reviews* |
| --- | --- | --- | --- | --- |
| Reviewer 1 | 36 | Top reviewers in Clinical Medicine - September 2018<br>Top reviewers for Medicine - September 2017 | Minerva Obstetrics and Gynecology (current); Guest Eeditor of 1 MDPI Special Issue | 253 |
| Reviewer 2 | 15 | none | Guest editor of 21 MDPI Special Issue | 0 |
| Reviewer 3 | 16 | none | Frontiers in Medicine (current); BMC Women's Health (past membership); Heliyon (current); Journal of Clinical Obstetrics & Gynecology (current); Technology in Cancer Research & Treatment (current) ; Open Journal of Obstetrics and Gynecology (current) Guest editor of 4 MDPI Special Issues | 99 |
| Reviewer 4 | 28 | none | Open Journal of Obstetrics and Gynecology (current); Guest editor of 2 MDPI Special Issues | 53 |
| Reviewer 5 | 42 | none | Guest editor of 54 MDPI Special Issues | 0 |
| Reviewer 6 | 15 | none | BMC Women's Health (current) | 0 |
| Reviewer 7 | 37 | none | BMC Women's Health (current); International Journal of Gynecology & Obstetrics (current); Journal of Biological Regulators and Homeostatic Agents (current); Journal of Ovarian Research (current); PLOS One (current); Guest editor of 4 MDPI Special Issues | 99 |
| Reviewer 8 | 34 | none | Reproductive Medicine | 15 |
| Reviewer 9 | 58 | Top Peer Reviewer Top reviewers in Biology and Biochemistry - September 2019; Top reviewers in Clinical Medicine - September 2019; Top reviewers in Cross-Field - September 2019; Top reviewers in Molecular Biology and Genetics - September 2019; Academy mentor; Excellent Reviewer (51) | Diagnostics (current); International Journal of Endocrinology (current); Journal of Clinical Medicine (current); Reproductive Medicine; Acta Chirurgica Belgica (current); Acta Endocrinologica (current); BMC Women's Health (current); Gynecologic and Obstetric Investigation (current); International Journal of Fertility & Sterility (current); Italian Journal of Gynaecology and Obstetrics (current); Journal of Minimally Invasive Gynecology (current); Journal of Ovarian Research (current); Minerva Surgery (current); Minimally Invasive Therapy and Allied Technologies (current); Neuroendocrinology (current); Open Medicine (current); Plos One (current); Scientific Reports (current); Translational Cancer Research (current); Wideochirurgia I Inne Techniki Maloinwazyjne (current); International Journal of Molecular Sciences (past); Experimental and Therapeutic Medicine (past); Guest editor of 20 MDPI Special Issues | 2,901 |

\* Peer review verified through Web of Science. It is possible that a scientist will have no 'verified peer reviews' even though they engage in peer review, if they have not performed the verification step through Web of Science.

## Discussion

We identified 195 review reports from a review mill, which largely consisted of boilerplate language, together with requests for citations to work authored by the review millers. As shown in Figure 2, the same citations were requested across multiple reviews by different individuals.

One might wonder whether a high rate of boilerplate comments is surprising: we selected review reports on the basis that they contained boilerplate comments, so we would expect to see such phrases. The key point to note, however, is that if a review was found by searching for one boilerplate comment, then it invariably contained many others. Furthermore, the same comments recurred not just within the corpus of review reports for one reviewer, but across reports from different reviewers, who were themselves co-authors of the other reviewers. This is evident by scanning the list of comments in online Supplementary Data 2. Indeed, if one sorts this list alphabetically by comment, large stretches of comments with identical wording are readily apparent. This indicates a level of co-ordination between reviewers.

The analysis of the comparison reviews established that the review mill reports were characterised not only by presence of boilerplate comments, but also by a low level of non-boilerplate comments. For many of these reviews, the only cases where a comment went beyond generic recommendations was to provide context for a suggested citation to the reviewer’s work.

Day (5) has suggested that “there are innocent reasons why referees might share template reports” and that presence of copied and pasted material across reviews does not necessarily indicate misconduct. However, an author deserves to have a peer review that engages with the content of the article; a few sentences that are duplicated across peer review reports may be acceptable but review reports that contain little or no original material engaging with the contents of a manuscript are not.

In sum, we argue that we have demonstrated beyond reasonable doubt the articles in our target set were not adequately scrutinized in the review process before they were incorporated in the corpus of academic literature. The network we identified corresponds to Oviedo-Garcia’s (2) definition of a review mill, and shows that this kind of research misconduct extends to clinical medicine. Review mills differ from other types of fake peer review, where the peer reviewer may be an illegitimate individual who operates under a false identity (6). Whereas fake peer review is often found as part of a paper mill (7), review mills do not involve the authors of the affected article.

We suspect the true number of articles affected by this review mill is much higher, given that there were over 2800 verified peer reviews by those we identified as review millers registered with Web of Science up until 7th October 2025 (see Table 3). This implies that we may have identified less than 10% of the peer review reports produced by this group of researchers.

Furthermore, 39% of the review mill reports were anonymous, so there could well be other members of the review mill that we cannot identify.

### Causes and consequences of review mills

The question arises as to what motivates review millers. In the case of the current review mill, we suspect that a goal is to increase citations and boost the reviewer’s H-index, which is widely regarded as an indicator of prestige and may lead to career advancement. We have shown that around 78% of articles complied with all or some of the requests for citations, leading individual reviewers to gain between 4 and 57 additional citations to their work. It is noteworthy that the review mill that we describe is based in Italy, a country where since 2011 government funding for universities as well as recruitment and promotion of professors is determined largely by bibliometric indicators based on publications and citations (8).

Another benefit for review millers would be to obtain discounts on Article Processing Charge (APC) for future works submitted to the same publisher: for instance, MDPI offers discount vouchers to selected reviewers to compensate them for their time and effort.

Compared to some other forms of academic malpractice, operating a review mill may seem a relatively minor peccadillo. Nevertheless, we argue that the practice should be taken seriously because of the impact review milling has on individuals and on the trustworthiness of science in general. In short, the articles that we identified have not undergone the kind of scrutiny that is expected from peer reviewers (9), making the authors of the affected papers victims of research misconduct. There is no reason to believe that original manuscripts that are reviewed by a review mill are flawed, but they are compromised by inauthentic peer review by reviewers who appear to use their position purely to suggest citations to their own work. In the case of another review mill, the publisher MDPI noted the problem with inadequately reviewed articles and proposed to re-review these (10); while this might help put the record straight, it can be devastating for authors to find the validity of their work questioned years after it was originally published. The publisher also suffers: having been proactive in promoting transparent peer review, they then are confronted with a host of questions on how to handle papers that were published without having met their peer review criteria. Will they be able to find new peer reviewers, and if so, do they inform them that the paper was previously published? What happens if a new peer reviewer recommends rejection or major revisions? Should the Article Processing Charge (APC) be refunded, and is the agreement between publisher and author still legally binding? Should there be a new status for the paper targeted by a review mill? What should happen to all the papers that have cited the affected work?

There is also a cost to other researchers. Academic research is a highly competitive field, and if some individuals obtain an unfair advantage by manipulating the peer review process, then other deserving individuals will lose out in terms of promotions and grants. The most serious consequences are for the scientific endeavour: the quality of published articles will be affected when review millers cynically treat peer review as an opportunity to boost their own citations, and offer only generic comments to authors. This is particularly serious in clinical medicine, where inadequately reviewed articles and unfairly boosted articles by reviewers may pollute meta-analyses and clinical protocols, as well as leading to potentially negative impacts on future research.

### Tackling review mills

In a similar way to false positive reviews, review mills are “a nightmare because they are difficult to recognize, require intensive investigations by the editor, and are suitable to corrupt the entire scientific publishing system” (6).We were able to document the phenomenon only because some publishers make peer review openly available. It would be dangerous to assume that review mills are confined to the journals we documented here: it is likely that they have infiltrated other publishers where they remain invisible and undetected. It could be argued that diligent editors should have been made suspicious by several indicators of manipulation of the peer review process as listed by the Committee on Publication Ethics (COPE) (11), including vague and generic reviews, requests to add self-citation, and reviewers who agree to review many manuscripts. However, if editors are involved in the review mill, then it may go undetected for years. We propose that to tackle this problem we need an open peer review process, ideally with identity of reviews and editors being available in meta-data for each article, together with the full text of peer reviews. Furthermore, if a review mill is detected, it is likely that a wide body of work is implicated, and other articles reviewed by the review millers should be checked.

We note that while publishers may be criticised for not defending better against review mills, the boilerplate reviews that we identified were not obviously fraudulent. When one sees large numbers of identical reviews, the problem is obvious (see Supplementary Data 2), but typically an editor would not be aware of anything amiss, especially since the identity of authors of the suggested citations is not evident unless the reference is checked. We were able to detect the review mill articles from publishers listed in Table 1 because these publishers followed best practice in using open peer review. It would be unfortunate if other publishers failed to adopt open practices for fear of being found to be affected by review mills. Of course, publishers have access to all peer reviews and indeed we are aware of growing interest by publishers in running automated plagiarism checks on reviews. However, if each publisher acts independently, there remains scope for review millers to go undetected if peer review is kept hidden. There is a strong case for the STM integrity hub (12) to facilitate sharing peer review reports between publishers, to help combat peer review mills.

### Conclusions

In conclusion, peer review milling threatens the scientific record and gives an unfair career advantage to members of the review mill. Whereas most cases of compromised publication integrity focus on authors (13), in the case of review mills, authors as well as publishers are victims. In medical contexts, patient safety is likely to be affected when clinical articles are inadequately scrutinized during peer review. We recommend that publishers adopt open peer review, employ automated checks, and transparently report the editors responsible for handling papers, to make detection of review mills easier for the entire scientific community.

## Data Availability Statement

The four supplementary data tables referred to in the text are available here: https://osf.io/k5acw/?view_only=4667f3c3b79a4566b38773f716ee967f. (This is a provisional link for peer review; a public link will be substituted for final version).

## Authors’ Contributions

**MAO**: Conceptualization, Data Curation, Methodology, Analysis, Writing. **RA**: Methodology, Analysis, Writing. **DB**: Data curation, Methodology, Visualization, Analysis, Writing.

## Ethics Committee approval

Not applicable.

